# A SCOPING REVIEW OF ALZHEIMER’S DISEASE HYPOTHESES: THE CASE FOR A MULTI-FACTORIAL THEORY

**DOI:** 10.1101/2023.07.26.23293030

**Authors:** Simon Duchesne, Louis-Simon Rousseau, Florence Belzile, Laurie-Ann Welsh, Beatrice Cournoyer, Marianne Arseneau, Véronick Lapierre, Sara-Maude Poulin, Olivier Potvin, Carol Hudon

**Affiliations:** Department of Radiology and Nuclear Medicine, Université Laval, Quebec City, QC, Canada; Quebec Heart and Lung Research Institute, Quebec City, QC, Canada; CERVO Brain Research Centre, Quebec City, QC, Canada; School of Psychology, Université Laval, Quebec City, QC, Canada; VITAM Research Centre, Quebec City, QC, Canada

**Keywords:** Alzheimer’s disease, theory, hypothesis, scoping review, amyloid, metabolism, mitochondrial dysfunction, cerebrovascular disease

## Abstract

**Introduction:** There is a common agreement that Alzheimer’s disease (AD) is inherently complex; otherwise, a general disagreement remains on its etiological underpinning, with numerous alternative hypotheses having been proposed.

**Methods:** We performed a scoping review of 105 original manuscripts describing hypotheses and theories of AD published in the past decades, characterized them as having a single or multifactorial focus, and tracked their impact.

**Results:** Three stages can be discerned in terms of hypotheses generation, with three quarter of studies proposing a hypothesis characterized as being single-focus. The most important theoretical groupings were the Amyloid group, followed by Metabolism and Mitochondrial dysfunction, then Cerebrovascular. Lately, evidence towards Genetics and especially Gut/Brain interactions came to the fore.

**Discussion:** When viewed together, these multi-faceted reports reinforce the notion that AD affects multiple sub-cellular, cellular, anatomical, and physiological systems at the same time but at varying degree between individuals. A major impediment remains provide a comprehensive view of all these systems and their interactions to manage its inherent complexity.

**RESEARCH IN CONTEXT:**

1. We propose a scoping review of 105 original manuscripts describing hypotheses and theories of Alzheimer’s disease (AD) that have been published in the past decades, characterized as having a single or multifactorial focus.
2. We found that three quarter of studies proposed a hypothesis characterized as being single-focus (77/105), with the most important theoretical groupings being the Amyloid group, followed by Metabolism and Mitochondrial dysfunction, then Cerebrovascular. Three stages can be discerned in terms of hypotheses generation. The first phase (∼1980-1995) included the establishment of the main thrusts that have endured to this day (Amyloid, Glial, Infection, Inflammation, Metabolism, Oxidative stress, and Proteinopathies hypotheses; multifactorality; and neurotoxicity). In the second phase (1995-2005), the importance of the Cerebrovasculature, Mitochondrial dysfunction, and Neurotransmitters were recognized. Lately (2005-2020), evidence towards Genetics (outside of the autosomal dominant form of AD), and especially Gut/Brain interactions came to the fore.
3. When viewed together, these multi-faceted reports reinforce the notion that AD affects multiple sub-cellular, cellular, anatomical, and physiological systems at the same time but at varying degree between individuals. A major impediment remains provide a comprehensive view of all these systems and their interactions to manage its inherent complexity.

**HIGHLIGHTS:**

- We propose a scoping review of hypotheses and theories for Alzheimer’s disease (AD)
- Out of over 11,000 abstracts, we reviewed 105 articles, separated as having a single-focus or multi-factorial approach
- The diversity of reports calls for an integrative view of AD in order to encompass its inherent complexity

## 1. INTRODUCTION

After more than a century of research, Alzheimer’s disease (**AD**) remains perplexing, a thorn in the side of modern medical science. Its etiological underpinning remains under debate, with numerous alternative hypotheses having been proposed. None were confirmed to a level sufficient for its operationalization into a therapeutic approach unequivocally proven effective and efficient in a clinical setting. Surveying this landscape of ideas – some vibrantly pursued, others nearly forgotten - should provide insights as to future general directions for research.

There is however a common agreement that AD is inherently complex. Its most common, pseudo-sporadic form is primarily associated with aging, and hence overlaps exist between the wide variability in anatomical and physiological changes that accompany the latter with those of the former, early in the disease process at the very least. Beyond aging but related to it, multiple risk factors have been identified from epidemiological studies, ranging from education in early life; hearing loss, traumatic brain injury, hypertension, alcohol consumption, and obesity at mid-life; to smoking, depression, social isolation, physical inactivity, air pollution, and diabetes, in later years (1). Yet, the compounding factor of time renders difficult the determination of whether some of these factors are early disease markers, etiological agents, catalysts, or a combination of these roles. The sheer breadth of these identified risk constructs, with impacts at scales ranging from proteins to behaviors, further points to the complexity of uncovering disease pathways at the most fundamental levels – and therefore amenable to therapeutic approaches, pharmacological or otherwise – that compound into the observed pathological cognitive and behavioral presentations. A common agreement on what constitutes AD seems therefore a first and necessary step to address this complexity and work together towards a solution. In fact, the discordance between the clinical presentation of AD and some of its most accepted biomarkers (e.g., amyloid beta (**Aβ**), present in AD but also prevalent at all ages without necessarily affecting cognition (2), and histopathological findings (e.g., similar lesions being associated with dementia or not (3)), means that the very definition of the disease remains a question as much of viewpoints as one of evidence. Thus, one could argue that there are three effective, overlapping yet still irreconciled definitions of AD: one arising from a clinical/cognitive viewpoint; a second, informed by *in vivo* biomarkers; and a third, defined by histopathological evidence.

An important aspect of this debate was introduced by Alois Alzheimer himself, when he first described the disease in his seminal paper (4), relating his observations on one patient, who had first come to the clinic at 51 years of age with clear clinical dementia. Based on the age of this patient and her rapid decline, it could be argued that this was a form of autosomal dominant AD. For the first time, Aβ plaques were observed and reported. Yet, this seminal observation has set the parameters of a major argument that still rages more than a century later: are Aβ plaques fundamental or accessory to the pseudo-sporadic form of the disease, form that is by far the most prevalent? Is AD an amyloidosis, first and foremost, in all cases – not only early in life, but in older patients as wellThis amyloid hypothesis (5), which has been at the center of research for the past few decades, is the field’s major attempt at a “great unifying theory”. It also oriented minds into a paradigm often seen in medical research, the search for a single etiological cause, regardless of the complexities inherent to human biology. Consequently, this singular view of the disease set in motion a quest, so far with limited positive results, for a similarly singular cure. Yet, the amyloid hypothesis is now seriously under siege. While it never made unanimity, as theories are initially wont to be, significant drawbacks in the past few years have led many to look anew for alternatives. First is the high prevalence of amyloid deposition in otherwise cognitively healthy individuals (6) and the lack of association between amyloid deposition and transverse and longitudinal cognitive status in otherwise cognitively healthy individuals (7). Next we can find the highly combinatorial nature of pathologies present (especially cerebrovascular lesions) when one looks at large post-mortem samples (8), meaning that amyloid is likely not the only aetiology driving decline. Finally, the accumulated failures of tens of phase III trials focused on Aβ removal to reach meaningful clinical changes (9), even when compared to those few that succeeded with modest (but, to be fair, statistically significant) success (10).

Thus, to solve the riddle of AD properly, one must find a theory able to explain these multiple events. Many have attempted to do so in the past. It is their work which is reviewed here (using the “Scoping Review” frame), in an attempt first to enumerate, then to assess the relative importance of the diversity of hypotheses and theories that have been proposed in the past decades. From this wide array of ideas, some conclusions are drawn to orient the field.

## 2. METHODS

### 2.1 Literature search strategy and sources

This review follows the PRISMA guidelines (11) and was conducted using the Web of Science, Embase, Medline, and PsycInfo databases for articles published from their inception through to 30 Oct 2020. Keywords referred to the variables of interest (AD theory). The search strategy is presented in Supplementary Material Table 1. Due to the sheer volume of work that has been published on AD, we limited the number of databases and search terms/synonyms. This qualifies our work as a scoping rather than systematic review.

### 2.2 Study selection process

Inclusion criteria were established as follows: (1) must be a peer-reviewed, scientific article; (2) must be written in either English or French; (3) must describe a hypothesis or theory related to AD; and (4) must not report solely on a pre-clinical model of AD .

Article selection was performed using the Covidence systematic review software (Veritas Health Innovation). After the initial search (L.S.R.), duplicates were removed. Next, a first sort was performed based on article titles and abstracts, followed by a second sort based on the full-text articles. All articles were evaluated against the inclusion and exclusion criteria, and some articles were excluded accordingly. The screening was made by independent reviewers (L.S.R.; F.B.; L.A.W.; B.C.; M.A.; V.L.; S.M.P.). Articles that were not included by reviewers were reevaluated by a third and included or excluded by common agreement.

### 2.3 Study selection

On 30 oct 2020, 11,023 studies were uploaded to Covidence from Embase (n = 3,142), Medline (n = 2,320), Web of Science (n = 4,240), and PsycInfo (n = 1,321) databases. Of these studies, 3,841 duplicates were removed. Two reviewers (L.S.R.; F.B.) screened the remaining 7,182 titles and abstracts according to the inclusion and exclusion criteria. The intervention of a third independent reviewer was not required. A total of 6,678 references were excluded. Full-text screening of the 504 articles led to the exclusion of 383 articles after review by six reviewers (L.S.R., L.A.W., B.C., M.A., V.L., S.M.P.). The excluded articles were either not describing a model (120), duplicates (86), described a non-etiological model (56), were proceedings (38) or non-peer reviewed articles (27), were in another language than English or French (23), were not applicable to AD (14), were inaccessible (12), were only concerned with the familial form of AD or Down syndrome (3), were not relevant for clinical (i.e. in humans) applications (2), or retracted (1)(see Figure 2).

**Figure 1.**
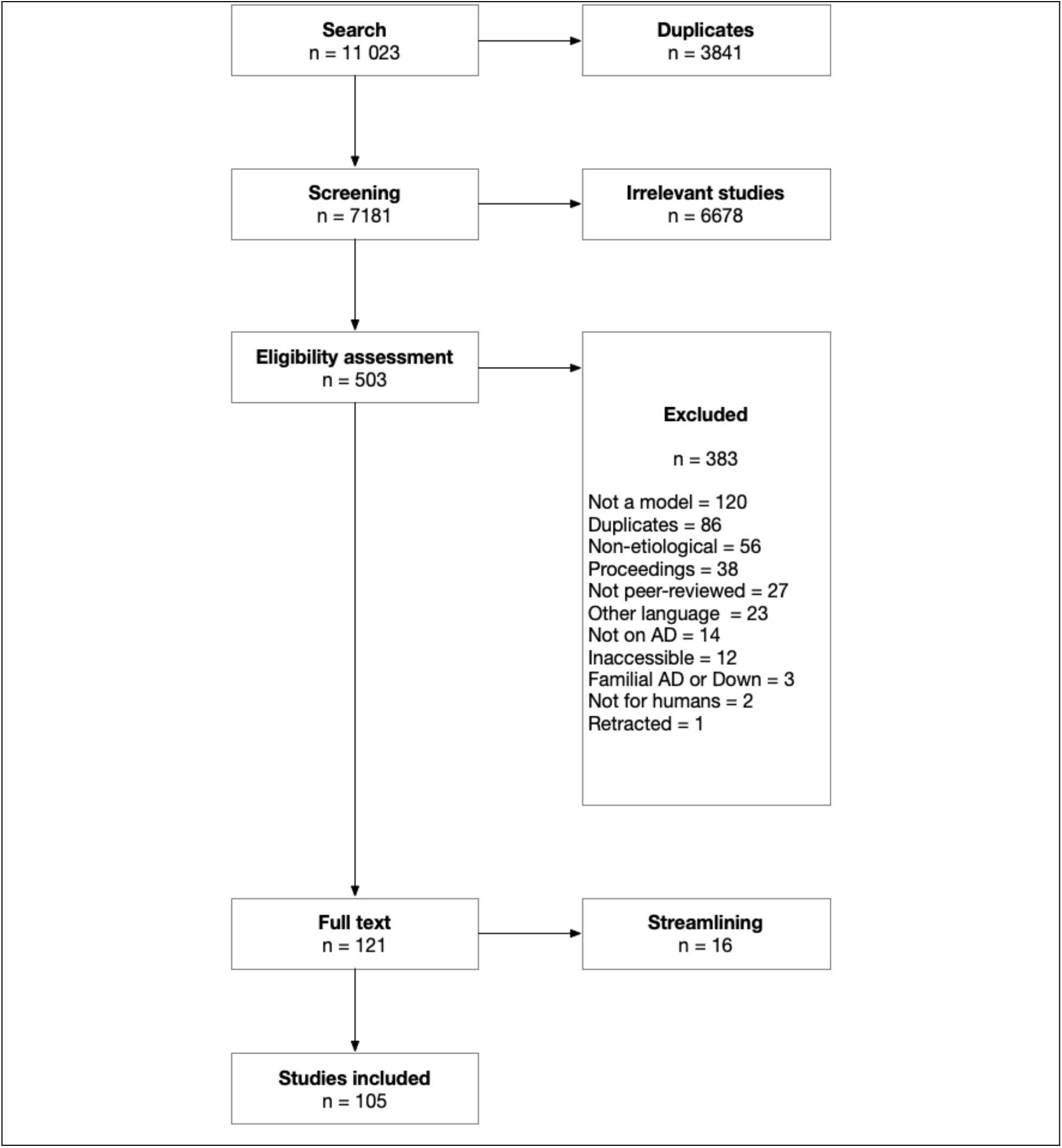
PRISMA flowchart. Following the search strategy, 11,023 abstracts were extracted from the Embase, Web of Science, Medline, and Psycinfo databases. After removing duplicates and irrelevant studies, applying exclusion criteria and streamlining to keep only seminal and original articles, a total of 105 studies were included in this scoping review.

**Figure 2.**
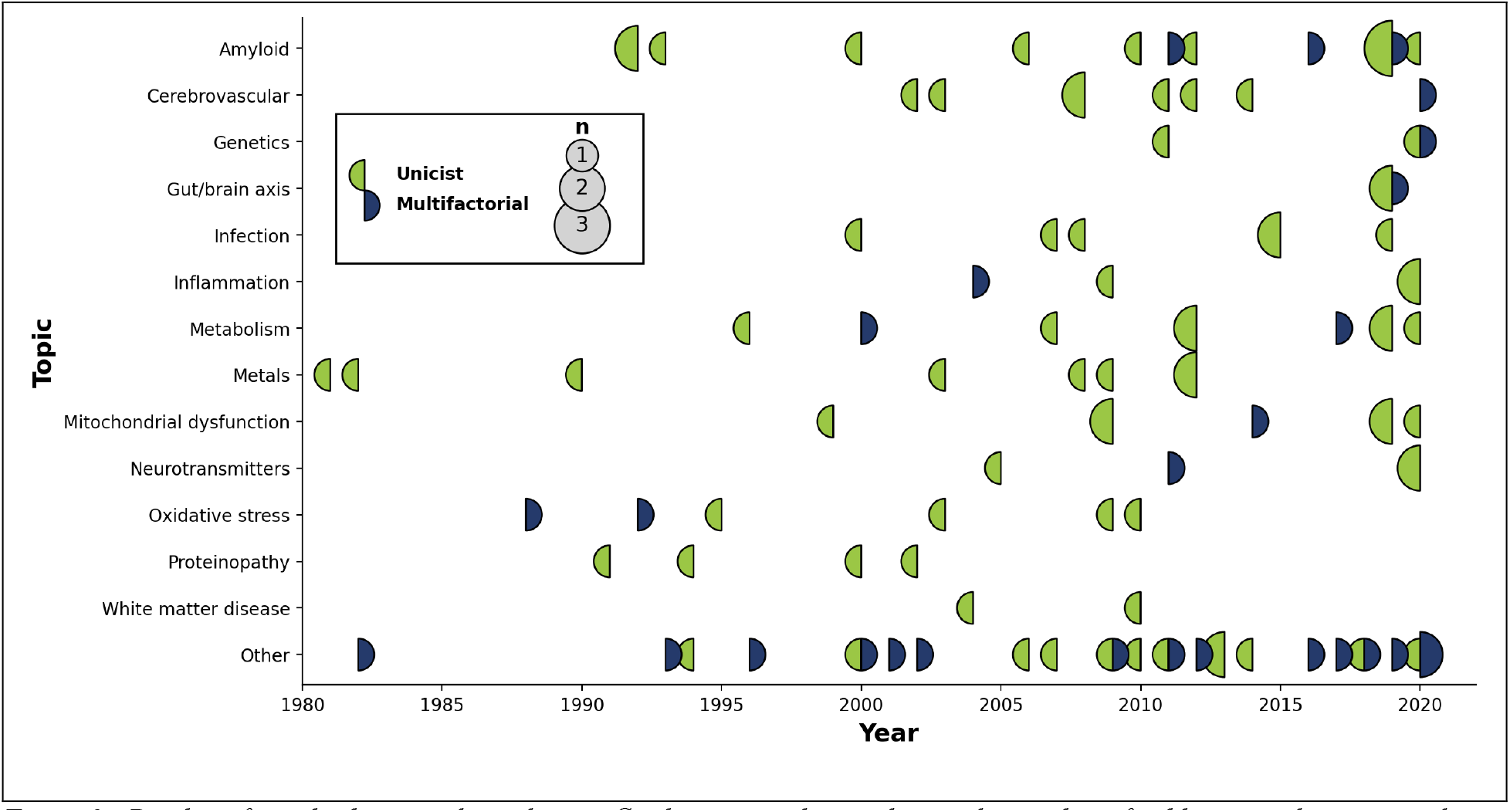
Display of articles by topic through time. Circles are sized according to the number of publications that year, with a color code indicating whether the published articles were unifactorial (left circle hemisphere) or multifactorial (right circle hemisphere)

A final streamlining review of the 121 remaining articles was completed (S.D.) to remove papers from an author or group of authors that published the same hypothesis/theory more than once. We included only the earliest paper in our review, as it was felt to represent the original inception of the idea. It is acknowledged that the latest manuscript, on the other hand, might have proposed the most up to date version of the hypothesis/theory under consideration.

Following this streamlining yielded a final tally of 105 papers that were included in this review (Supplementary Material Table 2).

### 2.4 Data extraction and classification

Characteristics of the included studies were extracted and are reported in Supplemental Material 2: lead author; country of lead author; whether there were other authors; title; year of publication; study design; and abstract.

Citations count for each paper as of 28 Feb 2023 were obtained via the Google Scholar search engine. While it is known that this count may include self-citations, it was the only count that could be reliably obtained for each paper across publishers and other citation engines.

Studies were then classified (S.D.) according to two epistemological dimensions. First was whether the hypothesis/theory being proposed was *unifactorial* (e.g., amyloid beta over-production initiates a cascade of downstream effects leading to neuronal death and dementia) or *multifactorial* (e.g., the co-existence and interactions between amyloid, tau, and inflammation lead to neuronal death and dementia). Secondly, studies were grouped according to the primary factor that was pushed forward in the article, according to the modified theoretical grouping of Ambrose 2015 (see Table 1).

**Table 1.** Theoretical groupings (Modified from Ambrose 2015)

| **Grouping** | **Description** | **Studies** |
| --- | --- | --- |
| Amyloid | Abnormal production, accumulation, or removal of amyloid-beta n-meric species and/or plaques | (12, 13, 14, 15, 16, 17, 18, 19, 20, 21, 22, 23, 24, 25) |
| Cerebrovascular | Vascular abnormalities affecting blood flow and perfusion, cerebrovascular lesions (infarcts, microbleeds, white matter hyperintensities) | (26, 27, 28, 29, 30, 31, 32, 33) |
| Genetics | Inherited genetic mutation(s), including APOE | (34, 35, 36) |
| Gut/brain axis | Dysbiosis in the gut microbiome | (37, 38, 39) |
| Infection | Protein misfolding in cells following infection by transmissible agents such as prion, subviral entities, HSV type 1 | (40, 41, 42, 43, 44, 45) |
| Inflammation/Immunological response | Autoimmune response and inflammation | (46, 47, 48, 49) |
| Metabolism | Cortical glucose utilization and transport across the blood-brain barrier | (50, 51, 52, 53, 54, 55, 56, 57, 58) |
| Metals | Neurotoxicity (e.g., Al, Hg) or deficiency in trace elements (e.g., Fe, Cu, Se) | (59, 60, 61, 62, 63, 64, 65, 66) |
| Mitochondrial dysfunction | Dysfunction in brain mitochondria leading to energy deficits and oxidative stress | (67, 68, 69, 70, 71, 72, 73) |
| Neurotransmitter | Deficits in cholinergic, noradrenergic, and other neurotransmitters | (74, 75, 76, 77) |
| Oxidative stress | Accumulation of oxidative damage in the brain from free radicals | (78, 79, 80, 81, 82, 83) |
| Proteinopathy | Tau, TDP43, and/or other proteins dysregulation | (84, 85, 86, 87) |
| White matter disease | Degeneration and disruption of white matter connections | (88, 89) |
| Others | Calcium channel blockers; nerve growth factors; autophagy, neuronal cell cycle abnormalities | (90, 91, 92, 93, 94, 95, 96, 97, 98, 99, 100, 101, 102, 103, 104, 105, 106, 107, 108, 109, 110, 111, 112, 113, 114) |

## 3 RESULTS

### 3.1 Studies overview

Out of 105 papers, a slim majority (58/105) was written by multiple, as opposed to a single, authors. First authors were mainly from the United States (51/105), Canada (7/105), or Europe (35/105) (see Table 2).

**Table 2.** First author provenance.

| Country | N papers |
| --- | --- |
| United States | 51 |
| Canada | 8 |
| France | 7 |
| UK | 6 |
| Italy | 6 |
| Australia | 4 |
| Germany | 3 |
| China | 2 |
| Poland | 2 |
| Spain | 2 |
| The Netherlands | 2 |
| Turkey | 2 |
| Brazil | 1 |
| Chile | 1 |
| Czech Republic | 1 |
| Hong-Kong | 1 |
| Iran | 1 |
| Israel | 1 |
| Portugal | 1 |
| Russia | 1 |
| Singapore | 1 |
| Switzerland | 1 |
| Total | 105 |

### 3.2 Study categories and impact

Three quarter of studies proposed a hypothesis characterized as being single-focus (77/105), with the most important theoretical grouping being, unsurprisingly, the Amyloid group (14/105), followed by Metabolism (9/105) (16/105 when joined to Mitochondrial dysfunction articles), and Cerebrovascular (8/105) (Table 3).

**Table 3.** Theoretical groupings frequency.

| **Grouping** | **N articles** | **N unifactorial** | **% unifactorial** | **Total citations** |
| --- | --- | --- | --- | --- |
| Amyloid | 14 | 10 | 71% | 9631 |
| Cerebrovascular | 8 | 7 | 88% | 904 |
| Genetics | 3 | 2 | 67% | 147 |
| Glia dysfunction | 2 | 2 | 100% | 441 |
| Gut/brain axis | 3 | 2 | 67% | 466 |
| Infection | 7 | 7 | 100% | 99 |
| Inflammation | 4 | 3 | 75% | 84 |
| Metabolism | 9 | 7 | 78% | 665 |
| Metals | 8 | 8 | 100% | 658 |
| Mitochondrial dysfunction | 7 | 6 | 86% | 1136 |
| Neurotransmitters | 4 | 3 | 75% | 576 |
| Oxidative stress | 6 | 4 | 67% | 985 |
| Proteinopathy | 4 | 4 | 100% | 153 |
| Other | 26 | 12 | 46% | 2109 |
| **Total** | **105** | **77** | **73%** | **18,054** |

Impact, as measured by total citations count, followed a similar pattern. Out of a total of 18,054 citations for the 105 papers, the most cited single-focused grouping was Amyloid (53.3%), followed by Mitochondrial dysfunction (6.3%, reaching 10.0% when adding Metabolism articles), then Oxidative Stress (5.5%) and Cerebrovascular (5.0%).

### 3.3 Evolution

The number of articles proposing, repositioning, or actualizing individual hypotheses and theories across time is shown in Figure 3, from the very first (King et al,’s aluminum toxicity hypothesis in 1981)(65) to the latest entries in 2020. Likewise, a similar timeline but this time showing the impact (i.e., citation count) for each one of these articles is shown in Figure 4.

**Figure 3.**
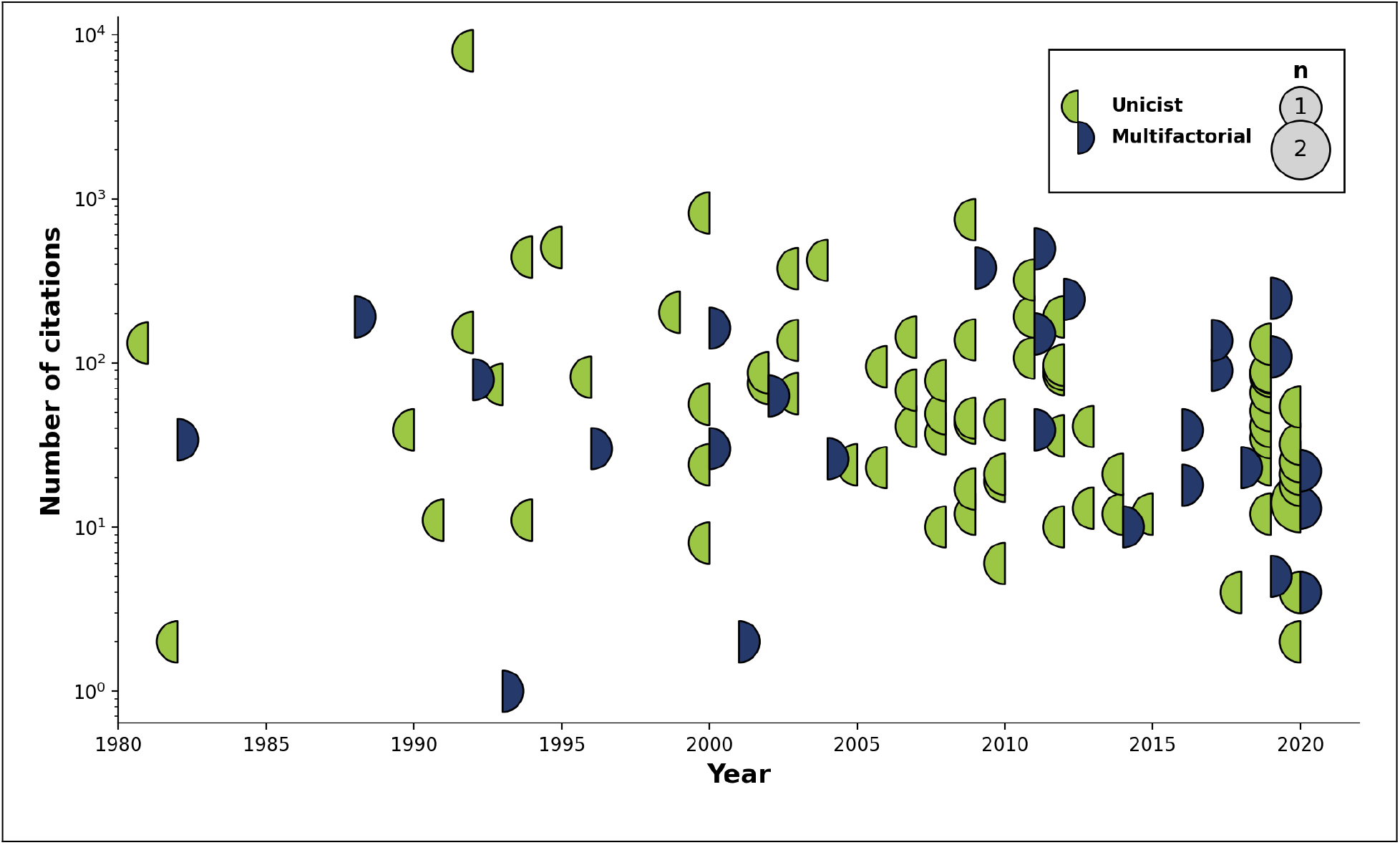
Display of articles’ citation count by topic through time. Circles are sized according to the number of citations that the articles achieved that year, with a color code indicating whether the published articles were unifactorial (left circle hemisphere) or multifactorial (right circle hemisphere)

## 4 DISCUSSION

### 4.1 Summary of findings

We have conducted a scoping review of AD-related hypothesis or theories in the literature dating from the early 1980s to the current era. We selected 105 papers for review, written mostly by authors based in North America and Europe, and spanning a wide range of conceptual groupings.

#### 4.1.1 Uni- or multi-factorial?

The idea that failure of a single pathway is responsible for the ensuing cascade of anatomical and physiological aberrations has been espoused not only for those hypothesizing about the role of Aβ, but all others as well, be they molecular, proteinic, cellular, or tissular entities, with roughly three quarters of all theories being unifactorial (*cf*. Table 3). Given the massive increase in neuroscientific knowledge of the last few decades, clearly adding to previous knowledge on the intricate complexity of the brain, and the lack of any clinically meaningful results in therapeutic pursuits when targeting individual pathways (*cf*. section 4.2), it remains puzzling why there remains such an emphasis on unifactorial hypotheses. This includes the Amyloid hypothesis subgroup, the most studied of them all, for which unifactoriality was *de rigueur* until 2011; since then, *more* unifactorial point of view articles have been published than those espousing a multifactorial framework in which Aβ is a strong contributor. In essence, the field seems to have doubled down on the amyloid cascade hypothesis, albeit modified, regardless of disproving evidence as to its centrality.

#### 4.1.2 Novelty and endurance of hypotheses

Three stages can be discerned in terms of hypotheses generation. The first phase (∼1980-1995) included the establishment of the main thrusts that have endured to this day. First was the Amyloid hypothesis, with the field-defining work of Hardy et al. (cited 7,991 times)(19), before the separation into clear autosomal dominant and pseudo-sporadic cases (often confounded with Early and Late Onset AD designations). In the same period were also introduced seminal reports for the Glial (16), Infection (44), Inflammation (81), Metabolism (55), Oxidative stress (81), and Proteinopathies (84) hypotheses; the concept of interdependence across all scales, central to multifactorality (104); as well as the by now marginal notion of neurotoxicity of trace elements (65). In the second phase (1995-2005), the importance of the Cerebrovasculature (33), Mitochondrial dysfunction (70), and Neurotransmitters (76) were recognized. Lately (2005-2020), evidence towards Genetics (outside of the autosomal dominant form)(36), and especially Gut/Brain interactions (37) came to the fore. The latter category has seen a rapid increase in impact when compared to others, with a rate of 62.0 citations/year since publication (second only to the seminal Hardy et al. paper, with 257.8 citations/year since publication).

#### 4.1.3 Biases

There are obvious biases in the impact statistics that must temper any interpretation. First, citation counts from Google Scholar often include self-citations; real numbers will therefore be smaller than recorded. Secondly, articles may be cited for positive (e.g., new evidence corroborating the hypothesis), negative (e.g., new evidence against the hypothesis), or even neutral (e.g., overview of the field) reasons, including positioning of one’s hypothesis against the leading one (which, in the case of AD, would favor the citation rates attached to the Amyloid group). Raw citation count therefore may not reflect actual acceptance of any idea. Finally, the field is heavily biased towards ground-breaking, first-in-field articles; while seminal, they may not represent the best or most up to date version of a particular hypothesis. In particular, the article of Hardy et al. (19) which we mentioned established the amyloid cascade hypothesis and was cited 7,991 times, represents 44.2% of *all* citations from the 105 papers in this scoping review. The power law of distributions is evidently quite at work here as elsewhere; the 10-most cited papers (Amyloid: 2; Cerebrovascular: 2; general theories around aging: 2; and Mitochondrial dysfunction, Oxidative stress, Neurotransmitters, and Glial disease: 1 each) are responsible for 69.2% of all citations.

### 4.2 Future considerations

#### 4.2.1 Which hypotheses or theory have notable evidence?

We posit that to answer this question successfully a theory would need to fulfill three criteria: 1) unequivocal clinical benefit from a therapeutic intervention, with 2) unambiguous disease-modifying results confirming a 3) global theoretical framework of the etiology.

As of now, only three classes of interventions have achieved results. First are anti-cholinesterase inhibitors, approved in the late 1990s (115) following clinical trials that demonstrated a statistically significant yet weak improvement in cognition for a limited time period. Acting on the principle that they enhance the availability of important neurotransmitters, they appear to provide but a temporary respite to clinical disease progression, while not influencing biomarkers of neurodegeneration. They remain seen as symptoms-, rather than disease-, modifiers (116). Hence, while our first criteria seem met, the latter two are not.

A second class of intervention has gained traction in the past few years. Three Aβ antibodies have been approved by the Food and Drugs Agency in the United States following successful clinical trials, for which the primary outcome was biomarker defined, rather than clinical (117). While the aptitude of both compounds at Aβ plaque removal is not in question (criteria 2), they had no effect on anatomy or physiology (e.g. neurodegeneration, neuroinflammation, neurometabolism) as well as limited clinically-observable effects (criteria 1), the latter remaining in doubt (9, 118), especially in the face of severe side-effects that point to an incomplete understanding of the role of Aβ (119).

Such antibodies are not the only compounds that have attempted to reduce production, block aggregation, or remove Aβ plaques; as a general rule, these few successes are not strong indicators of the centrality of the amyloid hypothesis in any general theory of AD (criteria 3).

Finally, non-pharmaceutical combinatory approaches targeting lifestyle interventions, nutrition, physical exercise, and aggressive management of co-morbidities (e.g. hypertension, diabetes) have had also significant successes, with effect sizes that are actually equal to pharmacological approaches such as Aβ antibodies (120)(criteria 1). The difficulty here is to understand which one, or many, etiological pathways are affected and with which efficiency (criteria 2). It does however provide strong evidence that *multi-scale targeting* stands a better chance than other, single-pathway focused interventions (criteria 3).

#### 4.2.2 Theory vs. practice: pharmaceutical trials

Hypothesis-driven research, relatively free from the pressure of providing immediate returns to its largely state funders in terms of success or failure, has therefore kept on exploring avenues, new and old, with what can be seem at times steadfastness. Witness to this effect the 33% *increase* in Aβ -centric theoretical works that have been proposed in the 2010-2020 interval.

On the contrary, the field of pharmaceutical trials seems to have already integrated the notion of multi-factoriality, exploring a host of different avenues, often in combination. This can be seen for example in the 54% *decrease* (41 vs. 19) in the number of Aβ-centric clinical trials between 2010 and 2022 (121) (122).

#### 4.2.3 Single vs. multiple factors

On the face of the evidence presented it is therefore hard to refute completely any one of the hypotheses that have been proposed. While many concepts can be proven as extremely unlikely and rejected on this basis, most articles present compelling evidence to this or that effect that is either borne out in a clinical population or animal model. Viewed differently, they reinforce the notion in fact that AD affects multiple sub-cellular, cellular, anatomical, and physiological systems at the same time but at varying degree between individuals. The biggest hurdle to overcome seems rather to provide a comprehensive view of all these systems at the same time, and their interactions. The emphasis on unifactorial viewpoints is possibly an artefact of research, including the well-known bias towards positive research reporting (123). The necessity to provide results that have reached statistical significance will naturally be easier to achieve in limited experimental settings, given statistical power considerations, and thus favour the study of individual effects over the logistically much more difficult task of deciphering interactions in a system; complexity increasing factorially with each new factor and parameter being considered.

## 4 CONCLUSION

Hence, while new frontiers keep being opened, it could be argued that most of the anatomical and physiological systems that impact or are impacted by AD, at various time and spatial scales, have already been well surveyed. A corollary of this observation is endurance. Indeed, some hypotheses have been proposed and therefore tested for over 30 years. The fact that we are still discussing whether AD is primarily an amyloidopathy, what is the importance of metabolism, or what role plays the cerebrovasculature, attests to both the complexity of the matter and the need for a holistic framework that could reframe the debate in simpler lines. Geometry provides an analogy: problems that are arduous in cartesian space become simpler when expressed in polar coordinates. Such a transformation might be necessary for the field to reach clarity.

## 5 ACKNOWLEDGMENTS AND SOURCES OF FUNDING

S.D. would like to acknowledge funding by the Canadian Institute for Health Research (Operating grant #PJT-159778). L.S.R. would like to acknowledge funding from the Fonds de recherche du Québec – Santé (scholarship #271206).

## 7 AUTHOR CONTRIBUTIONS

- Guarantors of integrity of entire study: all authors;
- Study concepts and design: S.D.; L.S.R.; C.H.;
- Literature research: L.S.R.
- Data acquisition and processing: L.S.R.; F.B.; L.A.W.; B.C.; M.A.; V.L.; S.M.P.; S.D.
- Methods, analysis and interpretation: L.S.R.; S.D.
- Manuscript preparation: S.D.; O.P.; C.H.
- Revision/review, all authors; and
- Manuscript definition of intellectual content, editing, and final version approval: all authors.

## Data Availability

All data produced in the present work are contained in the manuscript

