## Supplementary Material 1 for "A SCOPING REVIEW OF ALZHEIMER’S DISEASE HYPOTHESES: THE CASE FOR A MULTI-FACTORIAL THEORY"

**Search strategy –** **Pubmed**

(multifact*[tiab] OR multi-fact*[tiab] OR causa*[tiab] OR multimod*[tiab] OR multi-mod*[tiab] OR multidomai*[tiab] OR multi-domai*[tiab] OR multileve*[tiab] OR multi-leve*[tiab])

AND ("Models, Theoretical"[Mesh] OR hypothesi*[tiab] OR model*[tiab] OR theor*[tiab] OR framework*[tiab] OR frame-work*[tiab])

AND ("alzheimer disease"[MeSH Terms] OR alzheimer[tiab])

### **Search strategy - Embase**

(multifact* OR "multi-fact*" OR causa* OR multimod* OR "multi-mod*" OR multidomai* OR "multi-domai*" OR multileve* OR "multi-leve*"):ti,ab

AND ('theoretical model'/exp OR 'disease model'/exp OR 'biological model'/exp OR hypothesi*:ti,ab OR model*:ti,ab OR theor*:ti,ab OR framework*:ti,ab OR “frame-work*”:ti,ab)

AND ('Alzheimer disease'/exp OR alzheimer:ti,ab)

Cette recherche donne 1322 résultats.

### **Search strategy - PsycNet**

(multifact* OR multi-fact* OR causa* OR multimod* OR multi-mod* OR multidomai* OR multi-domai* OR multileve* OR multi-leve*)

AND ({Theoretical Interpretation} OR hypothesi* OR model* OR theor* OR framework* OR frame-work*)

AND ({Alzheimer's Disease} OR alzheimer)
